# A sex-specific genome-wide association study of blood lipid levels in All of Us

**DOI:** 10.1101/2025.11.21.25340771

**Authors:** Yunxi Li, In-Hee Lee, Sek Won Kong

## Abstract

Despite widely acknowledged sex differences in lipid metabolism and risks for cardiovascular disease, genetic associations contributing to such differences remain incompletely characterized. Here, we performed a sex-stratified genome-wide association study (GWAS) for four lipid profiles to identify loci exhibiting differential effects between males and females. Using whole-genome sequencing data from All of Us Research Program comprising 124,920 participants of diverse ancestry, we conducted GWAS analyses separately in males, females, and a pooled cohort. Our analyses validated previous findings on genes associated with lipid metabolism. In addition, we have found 5 genes showing significant sex-heterogeneous effects, including *CELSR2* showing stronger association in males for HDL-C (β*_female_*: -0.022, β*_male_*: -0.045); *GPAM* in females for HDL-C (β*_female_*: 0.042, β*_male_*: 0.013); *PLTP* in females for HDL-C (β*_female_*: 0.06, β*_male_*: 0.02); *ZPR1* in females for LDL-C (β*_female_*: 0.050, β*_male_*: 0.014); and *CMIP* in females for TG (β*_female_*: 0.037, β*_male_*: 0.019). These findings highlight sex-specific genetic contributions to lipid metabolism and underscore the importance of including sex in evaluating cardiovascular risk.

## Introduction

Studies have shown that there are sex-based differences in risk of heart disease. Ischaemic heart disease was found to occur 6-8 years earlier in males compared to females, potentially attributed to differences in lipid metabolism [1]. Females with heart failure with reduced ejection fraction are also found to have a lower risk of sudden cardiac death and arterial fibrillation, despite having more symptoms and depression compared to males [1]. In alignment with these findings, two cardiovascular risk-assessment tools suggested by the American Heart Association, Predicting Risk of CVD EVENTs (PREVENT) and Pooled Cohort Equations (PCE), both take sex into consideration and provide sex-specific risk assessments [2,3]. Key risk factors for cardiovascular disease include dietary risks, air pollution, tobacco, and high low density lipoprotein cholesterol (LDL-C) levels [4].

Serum concentrations of blood lipids are biomarkers for cardiovascular disease risk stratification [5]. Lipids are transported in the serum as lipoproteins, such as low density lipoprotein (LDL) and high density lipoprotein (HDL) [6]. LDL carries cholesterol to peripheral tissues via the bloodstream, where its accumulation within arterial walls promotes plaque formation, thereby contributing to increased cardiovascular risk [7]. One study using Mendelian randomization also showed associations between LDL-C and Alzheimer’s disease, inflammatory bowel disease, and type 2 diabetes [8]. HDL removes excessive cholesterol in peripheral cells through the reverse cholesterol transport (RCT) process, which contributes to HDL’s inverse relationship with cardiovascular risk [9,10]. In clinical blood panel measurements, LDL-C and HDL-C refer to the cholesterol content carried by these two lipoproteins [11]. In addition to LDL-C and HDL-C, concentrations of triglycerides (TG) and total cholesterol (TC) are also commonly measured in the blood lipid panel. TG serves as an efficient energy source and can be stored in adipose tissue [12]. Given its hydrophobic nature, TG is transported in triglyceride-rich lipoproteins like very low density lipoprotein (VLDL), and high levels of plasma TG is associated with increasing risk of atherosclerosis [13]. TC measures all types of plasma cholesterol and can also be used to evaluate cardiovascular risk [14].

Several genes and pathways have been found to affect lipoprotein and lipid metabolism in the liver and central nervous system (CNS). In the hepatic pathway, LDL receptors (*LDLR*) in the liver determines hepatic uptake of LDL, which in turn affects plasma LDL levels [6]. Apolipoprotein E (*APOE*) encodes a lipid transporter protein that transports lipids into cells through LDLR [15]. LDLR degradation is mediated by proprotein convertase subtilisin/kexin type 9 (PCSK9), which binds to LDLR and facilitates its degradation in lysosome [6]. For HDL, the synthesis process starts with binding cholesterol phospholipids from liver and intestine to ApoA-I (pre-beta HDL), facilitated by ATP-binding cassette transporter A1 (ABCA1)[6]. Lecithin-cholesterol acyltransferase (LCAT) facilitates the esterification of free cholesterol bound to HDL particles [6]. From there, HDL can be cleared by the liver through 2 pathways, either mediated by scavenger receptor class B type I (SR-BI) or cholesteryl ester transfer protein (CETP) [6]. Plasma TGs are removed from chylomicrons and VLDL the bloodstream through Lipoprotein lipase (LPL)-mediated lipolysis, where LPL is transported by lipoprotein-binding protein 1 (GPIHBP1) from endothelial cells into capillary lumen [16,17]. In the CNS, limited passage of cholesterol through the blood brain barrier (BBB) makes de novo cholesterol synthesis essential for brain development and brain homeostasis [18]. Cholesterol synthesis takes place mostly during the perinatal and adolescence ages, when myelination occurs on axons [18]. Despite adults having a more stable cholesterol level, cholesterol synthesis in astrocytes is still essential for growth and maintenance of neuronal components like myelin, synapse, dendrites, and axons [18].

Studies have shown that there are sex-based differences in lipid metabolism. More specifically, pre-menopausal women are found to have higher HDL-C and lower LDL-C levels compared to men [19,20]. One theory suggests that sex hormones mostly drive these sex-based differences, as observed in postmenstrual women and women with polycystic ovary syndrome (PCOS) on hormone replacement therapy, as well as transgender individuals on hormonal treatment [21]. In females, estrogen has been found to be associated with RCT process, more specifically with high levels of estrogen promoting hepatic uptake of cholesterol efflux from macrophages and higher ApoA-I synthesis rate, which is essential to HDL synthesis [20,22]. Lower LDL-C in females can potentially be attributed to increased apolipoprotein B-100 fractional catabolic rate [23]. Other studies have also questioned the sex-hormone theory explaining sex differences in lipid levels, considering that menopause and PCOS not only affects sex hormones but also involve body fat distribution and insulin sensitivity, which can be confounding factors when exploring sex hormone’s effect on lipid levels [19].

To date, no large-scale studies have rigorously examined the genetic contributions to male–female differences. A prior study, limited by insufficient sample size and modest effect estimates, yielded inconclusive findings that warranted further investigation [24]. Previous literature exploring genetic associations of lipid profiles are also limited by unequal representation of sex or did not explore sex-specific genetic associations [25,26]. In our study, we report a GWAS study on blood lipids based on the All of Us (AOU) cohort, one of the largest US population dataset with abundant electronic health record and genomic sequencing data. We aim to validate previously reported genetic loci and potentially find novel genetic loci associated with lipid metabolism. In addition, we aim to explore sex-specific genetic influence on lipid metabolism across age groups.

## Materials and methods

### Study population

AOU research program is an initiative led by the National Institute of Health that collects electronic health record and biosample data from 510+ sites across the United States [27]. Since its start of enrollment in 2018, the program has had 865,000+ participants as of July 2025, with a primary focus on participants aged from 18 to 85+ [27]. In our study, controlled tier data release version 8 was accessed in February 2025 [28].

We included participants with LDL-C, HDL-C, TG, Total Cholesterol, short read whole genome sequencing (srWGS) data, and sex assigned at birth. We included 4 concepts of measurements for LDL-C (with logical observation identifiers names and codes (LOINC) values 13457-7, 2089-1, 18262-6, 18261-8, and 49132-4), 3 concepts for HDL-C (LOINC values 2085-9, 49130-8, and 18263-4), 5 concepts for TG (LOINC values 2571-8, 3043-7, 3048-6, 12951-0, and 1644-4), and 1 concept for TC (LOINC 2093-3). Measurements with missing measurement value or unit were removed. Measurements using operators <, ≤, >, ≥ or units other than mg/dL were also excluded from analysis. We did not convert other reported units to mg/dL because the values would be extremely high after conversion, indicating that there are likely some manual entry errors of units. Negative LDL-C values were excluded. For cholesterol measurements, we selected the highest LDL-C value for each person, and retained the participants with HDL-C, TG, and Total Cholesterol measurements available at exactly the same timepoint as LDL-C (Figure 1). A total of 124,920 participants were included in the analysis, comprising 75,127 (60.1%) females and 49,793 (39.9%) males.

**Figure 1.**
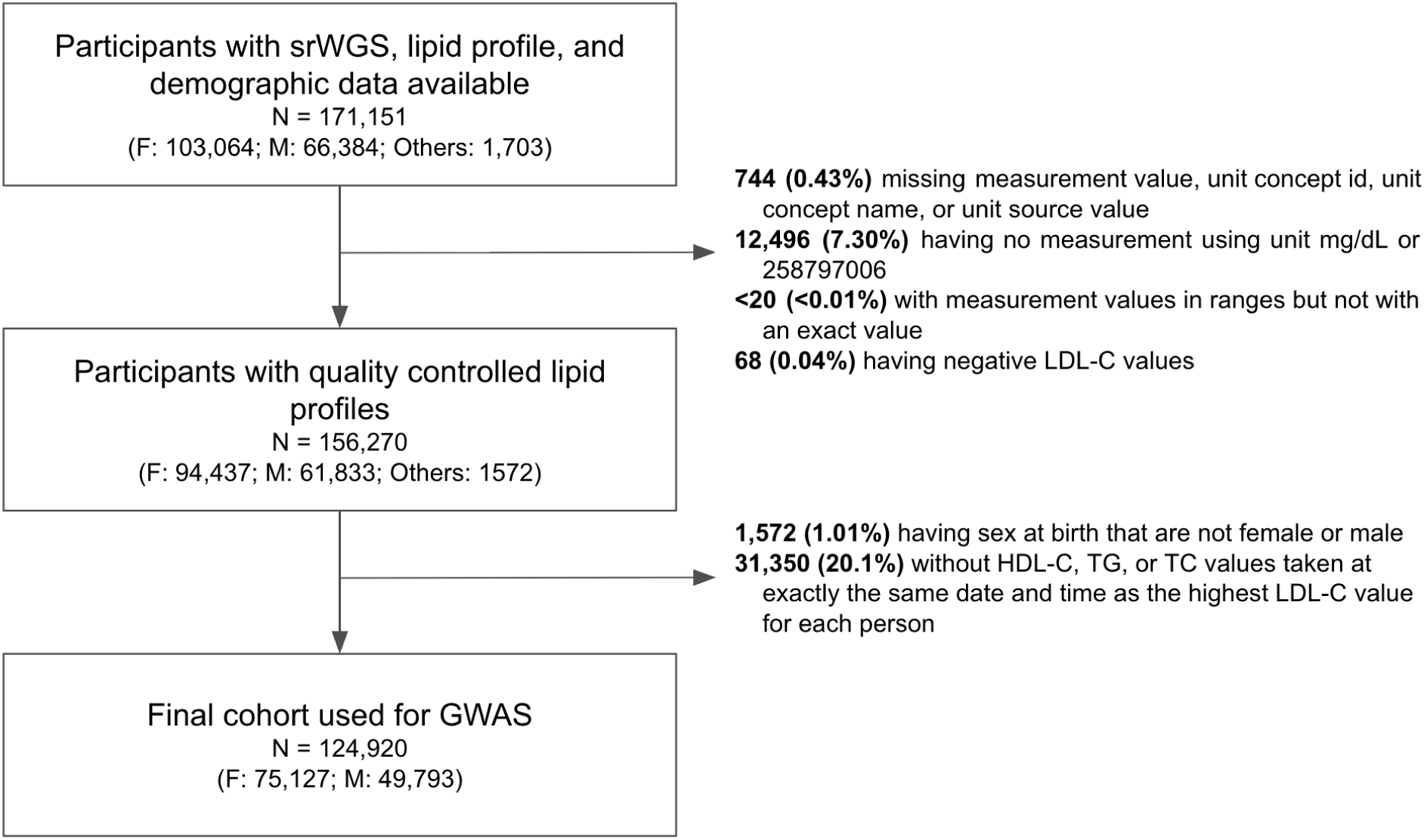
Flowchart of cohort selection. The figure outlines the sequential filtering of participants with whole-genome sequencing data, lipid measurements, and demographic information. After removing individuals with incomplete or invalid lipid values and those without complete paired lipid measurements, a final cohort of 124,920 participants (75,127 female; 49,793 male) was retained for genome-wide association analysis (GWAS) (srWGS: short-read whole genome sequencing data; HDL-C: high density lipoprotein cholesterol; LDL-C: low density lipoprotein cholesterol; TG: triglyceride; TC: total cholesterol; mg/dL: milligrams per deciliter; Others: participants whose sex-at-birth field contained “I prefer not to answer,” “pmi: skip,” “sex at birth none of these,” or “intersex”).

Statin usage history within 1 year before the time of measurement was extracted. Given the extensive missingness of end time of statin use records and variable impact of statin withdrawal on lipid profiles, it is not feasible to comprehensively analyze if lipid profile taken under a specific timepoint was under the influence of statin [29]. However, given that the highest LDL-C measurement was selected for each participant followed by matching HDL-C, TG, and TC measured at the same time, the measurements are highly likely to not be affected by statin treatment and most likely due to genetic and dietary factors. Obesity history within 1 year before the time of measurement was extracted. For body mass index (BMI) measurements, the BMI measurement closest to time of cholesterol measurement was extracted for each participant. Genetic ancestry prediction was extracted from the AOU data release v8 [30].

### Genome-wide association analysis

PLINK2 was used for quality control (QC) of srWGS data prior to pooled and followed by sex-specific GWAS [31]. We followed REGENIE’s quality control steps on UK Biobank data, which included filtering out variants with a minor allele frequency (MAF) ≤ 1%, Hardy-Weinberg disequilibrium (HWE) p-value <1 × 10^−15^, and a genotyping rate < 99%. We performed linkage disequilibrium (LD) pruning using a sliding window of 1,000 markers, a step size of 100 markers, and an r² threshold of 0.9 [32]. After QC, there are 1,441,940 SNPs to be tested in pooled GWAS, 2,739,534 to be tested in female-specific GWAS, and 2,971,581 to be tested in male-specific GWAS.

We used REGENIE v2.0.2 for pooled and followed by sex-specific whole-genome-wide association study. We selected REGENIE for its capability of handling large scale biobank data and conducting analysis of multiple phenotypes in parallel [26,32]. Rank based inverse normal transformation was performed for each lipid type to normalize lipid panel data [32,33]. In this sense, each unit change in effect size of REGENIE output was predicting inverse normal transformed lipid panel values. Age and first 16 genetic principal components were included as covariates for all GWAS runs. For pooled analysis, sex was additionally included as a covariate. A genome-wide significance threshold of *P* < 5 × 10⁻⁸ was used to determine significant SNPs to account for multiple testing of four lipid types [34]. We also applied Bonferroni correction to account for multiple testing of 4 blood lipid profiles. Beta coefficients were estimated using the reference allele as the effect allele. Regional association plots were created using the locuszoomr package [35].

### Gene-based analysis

REGENIE tested SNPs were converted to GRCh37 to use FUMA to detect genomic risk loci for each lipid profile and sex [36]. Genomic risk loci were defined as independent significant SNPs in summary statistics and surrounding SNPs based on linkage disequilibrium (LD) structure [36]. Region plots for each genomic risk loci were plotted for both pooled and sex-specific GWAS. The nearest gene for each leading SNP was annotated by MAGMA, where the most deleterious annotation is annotated for a region of multiple overlapping genes, and more than 1 gene can be annotated in the case of intergenic regions [36,37]. Leading SNPs across genomic risk loci were cross-referenced with previously reported lipid-associated loci by comparison against the GWAS Catalog (All Associations v1.0.2), using rsID as the identifier [38].

### Sex-stratified genotype effects on lipid levels

To evaluate genotype effects on lipid levels, we fitted separate age-adjusted linear models for males, females, and the pooled cohort. Within each sex stratum, individuals were grouped according to their genotype (reference homozygote, heterozygote, or alternate homozygote). For each subset, we fitted an analysis of covariance (ANCOVA) model of the form: Value ∼ genotype + age. Estimated marginal means (EMMeans) were derived using the emmeans package in R to obtain adjusted mean lipid levels for each genotype after controlling for age. Pairwise genotype contrasts were evaluated using Tukey’s significance adjustment to account for multiple comparisons. Analyses were performed separately for each sex and for the combined cohort.

## Results

### Study cohort and distribution of lipid profiles in female and male

The total cohort size was N = 124,920 including 75,127 (60.1%) female and 49,793 (30.9%) male. The majority of participants were of European ancestry (67.1%), followed by individuals of African ancestry (17.3%). The proportions of participants who had statin use history within 1 year before the date of lipid profile measurement were 13.8% (female) and 22.3% (male).

Females had higher mean LDL-C, HDL-C, and TC levels, while males had higher TG levels (Table 1). When stratified by age, females and males exhibited distinct age-related distributions of blood lipid levels (Figure 2). Females have lower median LDL-C levels in early ages (before age 50). However, females have higher median LDL-C levels after menopause, surpassing males of the same age group. This is consistent with previous findings on females having higher LDL-C from early adulthood to middle-age when compared with males [39]. Male, on the other hand, have higher TG levels than females, especially in early ages when compared to females in the same age groups. This is consistent with previous finding that males have a more atherogenic lipid profile [39]. Total Cholesterol levels for both sexes are similar in early ages, but females have higher total cholesterol levels after menopause (around age 50). This can potentially be attributed to the change in LDL-C levels as age increases. Across all age groups, females have higher median HDL-C compared to males, which is consistent with previous review literature [39].

**Figure 2.**
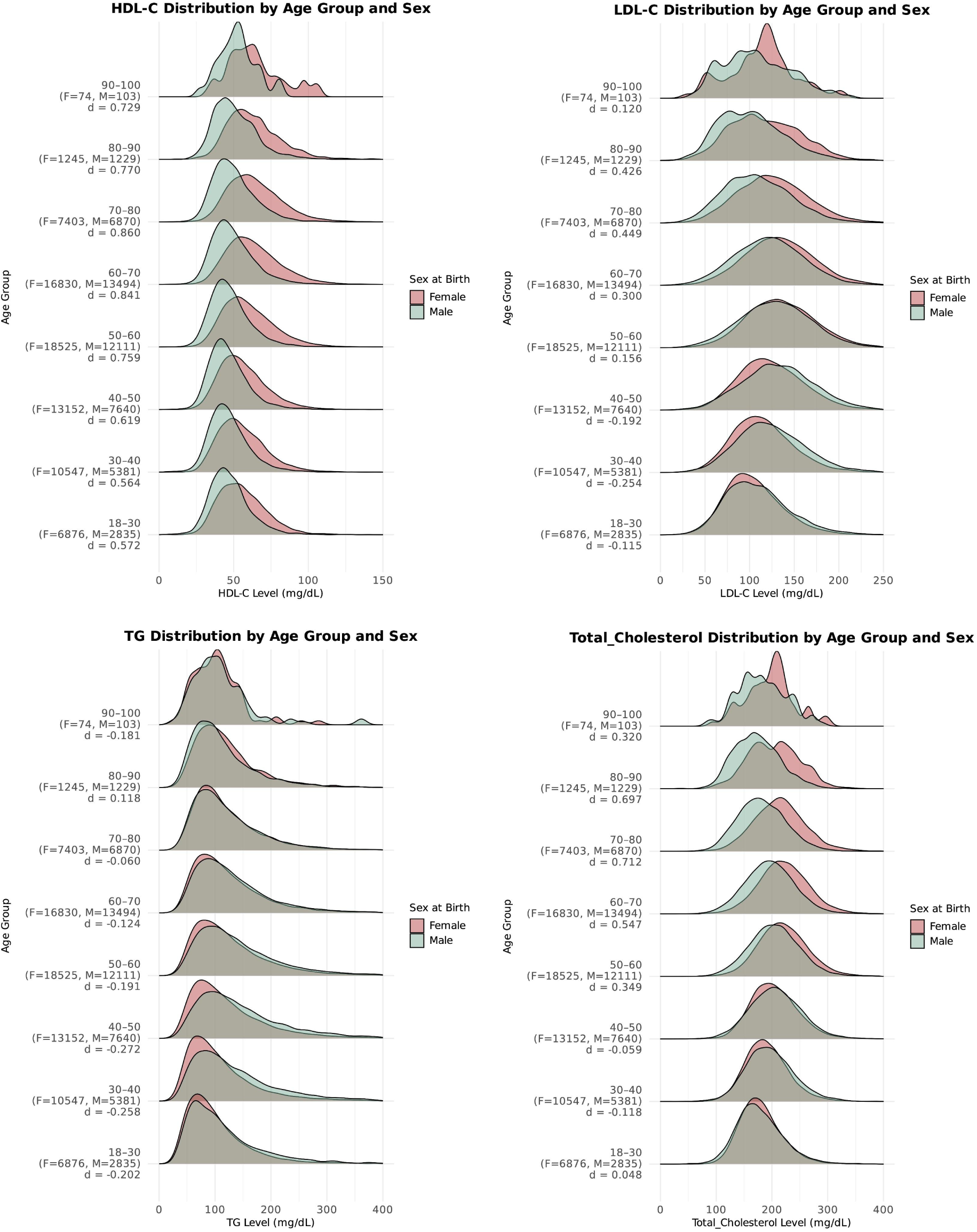
Age and sex differential distribution of HDL-C, LDL-C, TG, and Total Cholesterol. Density plots show HDL-C and LDL-C distributions across age groups for females and males. For each age stratum, sample sizes and Cohen’s d are reported where Cohen’s d quantifies the standardized mean difference between sexes using the following equation (positive d means higher mean in females).

**Table 1.**
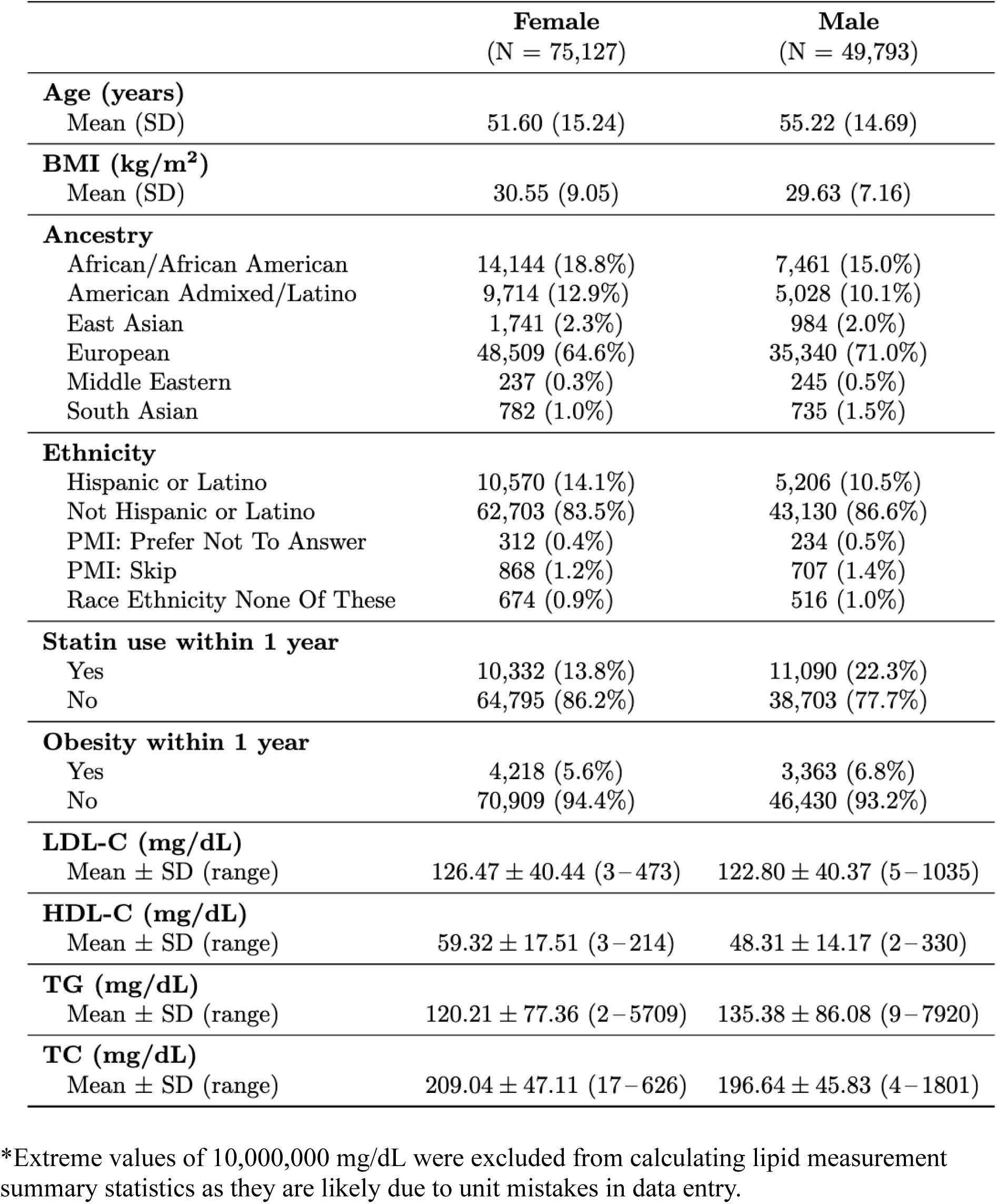
Baseline characteristics of study cohort.

### Genome-wide association analysis

We identified 1,766 genome-wide significant variants after Boneferroni correction taking into account multiple testing across 4 lipid types, including 541 for HDL-C, 384 for LDL-C, 445 for TG, and 396 for TC (Supplementary Table S1).

LDL-C and TC did not show strong genetic inflation across all 3 GWAS, while HDL and TG showed higher genetic inflation (Supplementary Figure 4). We did not find novel loci compared to previously reported analysis on GWAS catalog as indexed in August 2025.

Gene-based association analysis using MAGMA identified 298 statistically significant genes after applying Bonferroni correction for multiple testing across all tested genes (corrected p < 0.05), including 73 for HDL-C, 72 for LDL-C, 84 for TG, and 69 for TC (Supplementary Table S2). Our results validated previous findings of genes associated with lipid profiles as reported on GWAS catalog and we did not find novel loci associated with lipid profiles. We validated *LPL*, *ABCA1*, *CETP* to be associated with HDL-C; *PCSK9*, *CELSR2*, *LDLR*, and *APOE* to be associated with LDL-C; *LPL* to be associated with TG (Figure 3) [40].

**Figure 3.** Manhattan plots for pooled GWAS. Manhattan plots display pooled genome-wide association results for HDL-C (Figure 3A), LDL-C (Figure 3B), TG (Figure 3C), and TC (Figure 3D) in the pooled cohort. Each point represents a genetic variant plotted by chromosomal position and –log₁₀(P). The horizontal dashed line denotes the genome-wide significance threshold after Bonferroni correction.

**Figure 3A.**
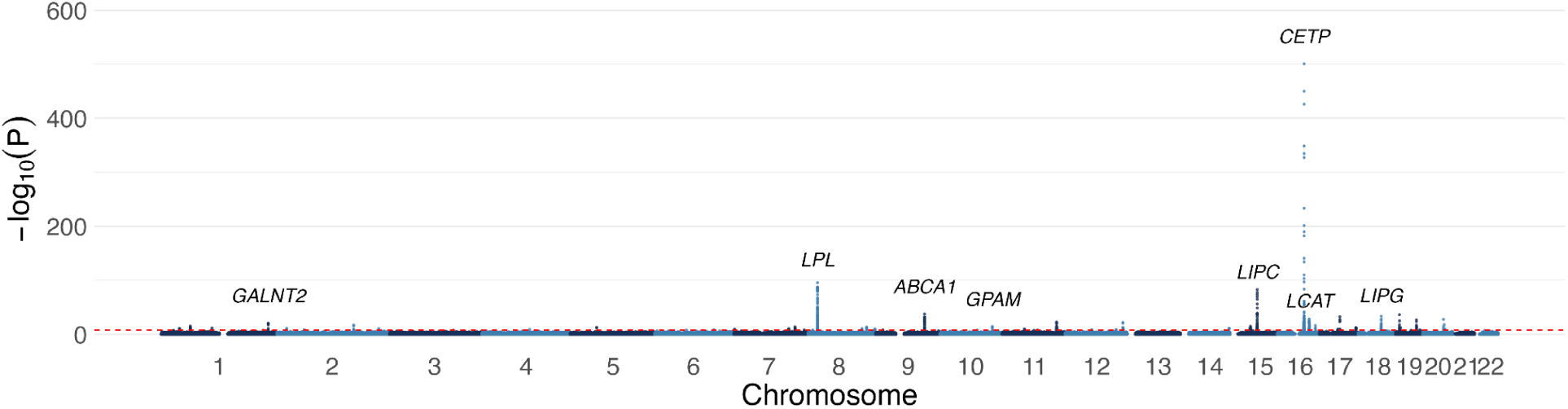
Manhattan plots for pooled GWAS for HDL-C.

**Figure 3B.**
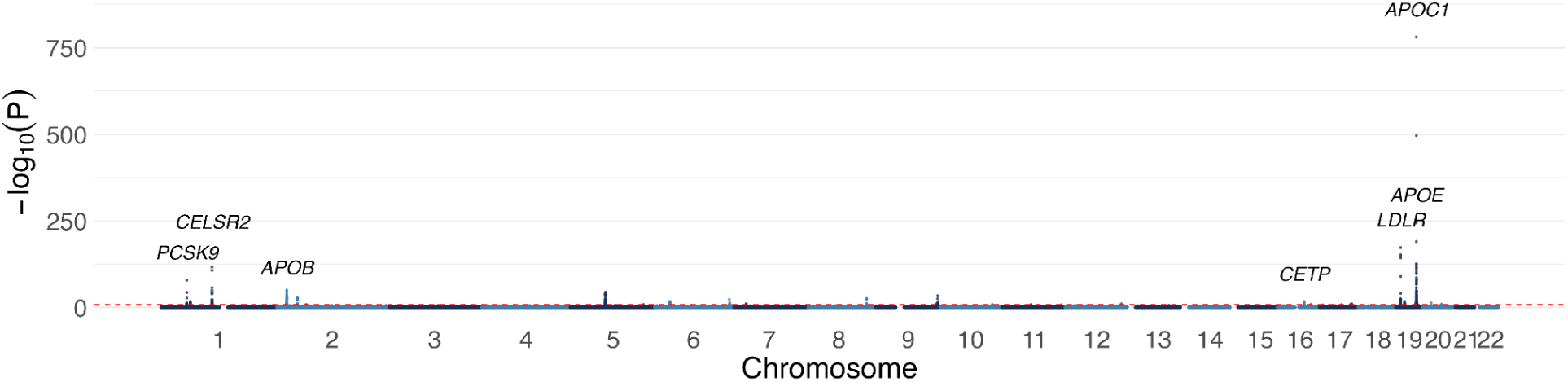
Manhattan plots for pooled GWAS for LDL-C.

**Figure 3C.**
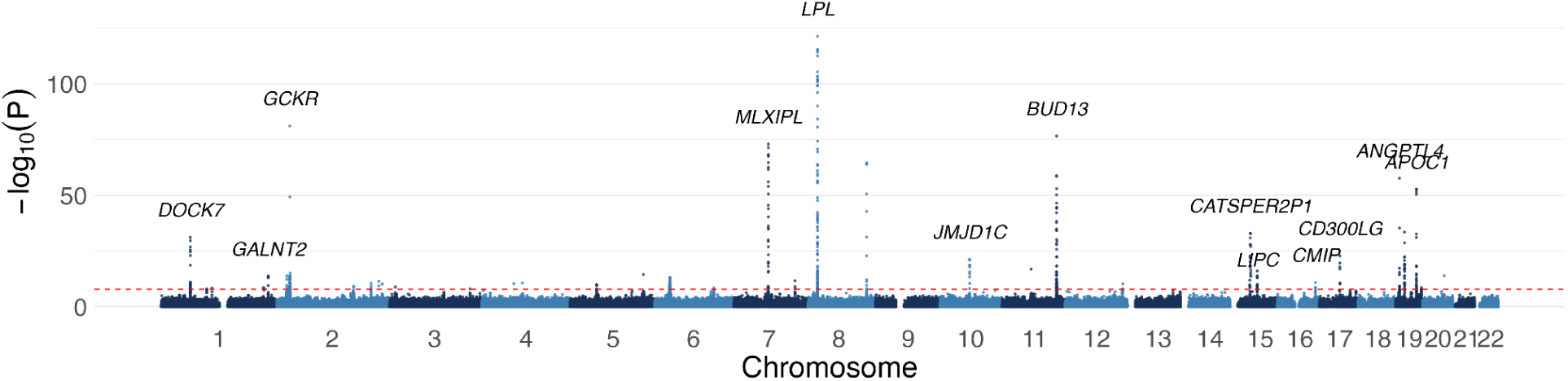
Manhattan plots for pooled GWAS for TG.

**Figure 3D.**
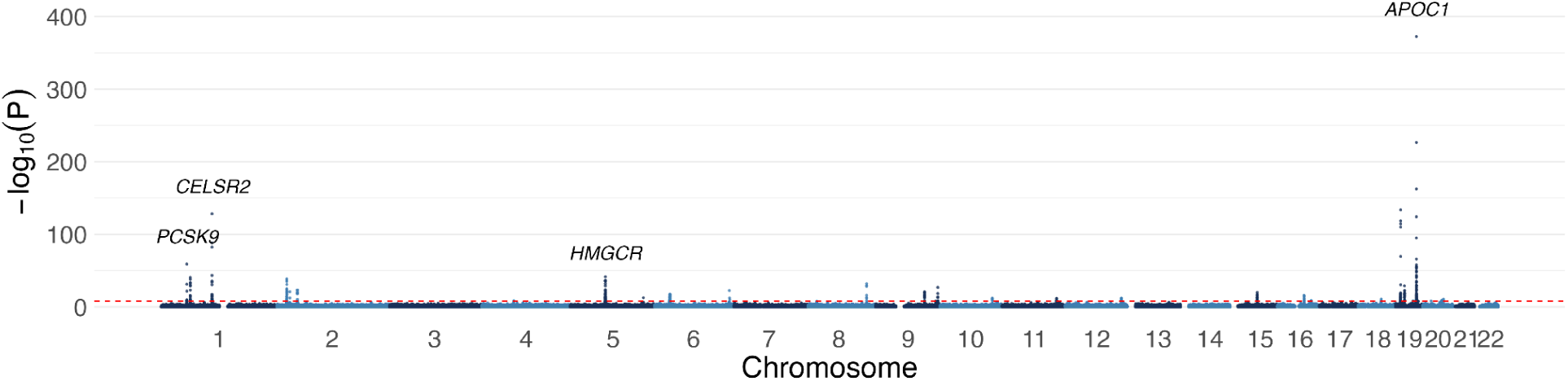
Manhattan plots for pooled GWAS for TC.

### Sex-stratified genome-wide association analysis

Following the pooled GWAS analysis, we performed sex stratified GWAS analysis for all 4 blood lipids (Supplementary Figure 1). In female-specific GWAS, we identified 2,227 genome-wide significant variants after Boneferroni correction taking into account multiple testing across 4 lipid types, including 568 for HDL-C, 506 for LDL-C, 593 for TG, and 560 for TC (Supplementary Table S3). In male-specific GWAS, we identified 1306 genome-wide significant variants, including 430 for HDL-C, 231 for LDL-C, 455 for TG, and 190 for TC (Supplementary Table S4).

Gene-based association analysis using MAGMA identified 267 statistically significant genes for female-specific GWAS run after applying Bonferroni correction for multiple testing across all tested genes (corrected p < 0.05), including 53 for HDL-C, 68 for LDL-C, 69 for TG, and 77 for TC (Supplementary Table S5). For male-specific GWAS run, we found 131 statistically significant genes, including 34 for HDL-C, 30 for LDL-C, 44 for TG, and 23 for TC (Supplementary Table S6). Manual review for removing spurious associations and identifying sex-specific associations further refined the list to 4 loci for HDL-C, 2 loci for LDL-C, and 1 loci for TG (Table 2). Among these sex-specific loci, *CELSR2* was male-specific for HDL-C, and all others were female-specific. Variant level and gene level tables are shown in Table 3 and Table 4, respectively.

**Table 2.** Sex-specific genes associated with blood lipid levels.

|  | **HDL-C** | **LDL-C** | **TG** | **TC** |
| --- | --- | --- | --- | --- |
| **Male** | *CELSR2* | - | - | - |
| **Female** | *GPAM, PLTP* | *ZPR1* | *CMIP* | - |

**Table 3.** Sex-specific variants associated with blood lipid levels.

| **ID** | **VEP_mapped_gene** | **BETA\*** | **P** | **PHENOTYPE** | **GWAS Run** |
| --- | --- | --- | --- | --- | --- |
| chr1:109274570:A:G | *CELSR2* | -0.0220579 | 0.000130701 | HDL | Female-only |
| chr1:109274570:A:G | *CELSR2* | -0.0448627 | 1.92E-10 | HDL | Male-only |
| chr1:109274570:A:G | *CELSR2* | -0.0314057 | 2.36E-12 | HDL | Pooled |
| chr10:112150963:A:G | *GPAM* | 0.0321485 | 3.02E-15 | HDL | Pooled |
| chr10:112153464:T:C | *GPAM* | 0.0422569 | 3.34E-16 | HDL | Female-only |
| chr10:112153464:T:C | *GPAM* | 0.0129896 | 0.0392735 | HDL | Male-only |
| chr11:116778201:G:C | *ZPR1* | 0.0494462 | 7.68E-14 | LDL | Female-only |
| chr11:116778201:G:C | *ZPR1* | 0.0140275 | 0.105052 | LDL | Male-only |
| chr16:81486161:A:G | *CMIP* | 0.0373586 | 2.74E-11 | TG | Female-only |
| chr16:81486161:A:G | *CMIP* | 0.0186053 | 0.00833067 | TG | Male-only |
| chr16:81486161:A:G | *CMIP* | 0.0297154 | 1.49E-11 | TG | Pooled |
| chr20:45916308:T:C | *PLTP* | 0.0562298 | 4.39E-19 | HDL | Female-only |
| chr20:45916308:T:C | *PLTP* | 0.0240854 | 0.00163817 | HDL | Male-only |
| chr20:45916308:T:C | *PLTP* | 0.0429295 | 1.43E-18 | HDL | Pooled |

\*Beta coefficients were estimated using the reference allele as the effect allele.

**Table 4.** Sex-specific genes associated with blood lipid levels.

| Gene | Locus | P | NSNPs | Phenot<br>ype | GWAS Run |
| --- | --- | --- | --- | --- | --- |
| *CELSR2* | chr1:109249539-109275751 | 0.00062272 | 36 | HDL-C | Female-only |
| *CELSR2* | chr1:109249539-109275751 | 4.87E-07 | 40 | HDL-C | Male-only |
| *CELSR2* | chr1:109249539-109275751 | 1.65E-09 | 23 | HDL-C | Pooled |
| *GPAM* | chr10:112149865-112215416 | 1.21E-13 | 50 | HDL-C | Female-only |
| *GPAM* | chr10:112149865-112215416 | 0.20378 | 54 | HDL-C | Male-only |
| *GPAM* | chr10:112149865-112215416 | 4.54E-10 | 36 | HDL-C | Pooled |
| *PLTP* | chr20:45898621-45912235 | 3.33E-15 | 18 | HDL-C | Female-only |
| *PLTP* | chr20:45898621-45912235 | 0.00033633 | 19 | HDL-C | Male-only |
| *PLTP* | chr20:45898621-45912235 | 0.0068117 | 9 | HDL-C | Pooled |
| *ZPR1* | chr11:116773799-116788046 | 2.27E-12 | 9 | LDL-C | Female-only |
| *ZPR1* | chr11:116773799-116788046 | 0.44892 | 11 | LDL-C | Male-only |
| *ZPR1* | chr11:116773799-116788046 | 0.48202 | 2 | LDL-C | Pooled |
| *CMIP* | chr16:81444808-81711762 | 4.20E-15 | 518 | TG | Female-only |
| *CMIP* | chr16:81444808-81711762 | 0.080907 | 575 | TG | Male-only |
| *CMIP* | chr16:81444808-81711762 | 9.40E-11 | 291 | TG | Pooled |

Examination of the regional association plot surrounding *CELSR2* region revealed clusters of SNPs in strong linkage disequilibrium exhibiting genome-wide significance after Bonferroni correction in the male and pooled cohorts, but not in females (Figure 4A). We observed a male-specific dosage effect of increasing HDL-C of variant chr1:109274570:A:G (rs7528419) (Figure 5). EMMeans derived from age-adjusted models revealed genotype-dependent differences in HDL-C levels in males but not females. In the male-only cohort, individuals carrying one or two alternate alleles exhibited progressively higher adjusted means compared to reference homozygotes (Tukey-adjusted *p* < 0.05). Such an effect was not observed in the female-only cohort.

**Figure 4.** Regional association plots showing sex-specific associations. Regional association plots illustrate sex-specific associations. Variants are shown with –log₁₀(P) values plotted against genomic position, and are colored according to linkage disequilibrium (r²) with the lead variant. Gene models and transcript orientations are displayed below each plot to contextualize the association signals using the locus zoom package.

**Figure 4A.**
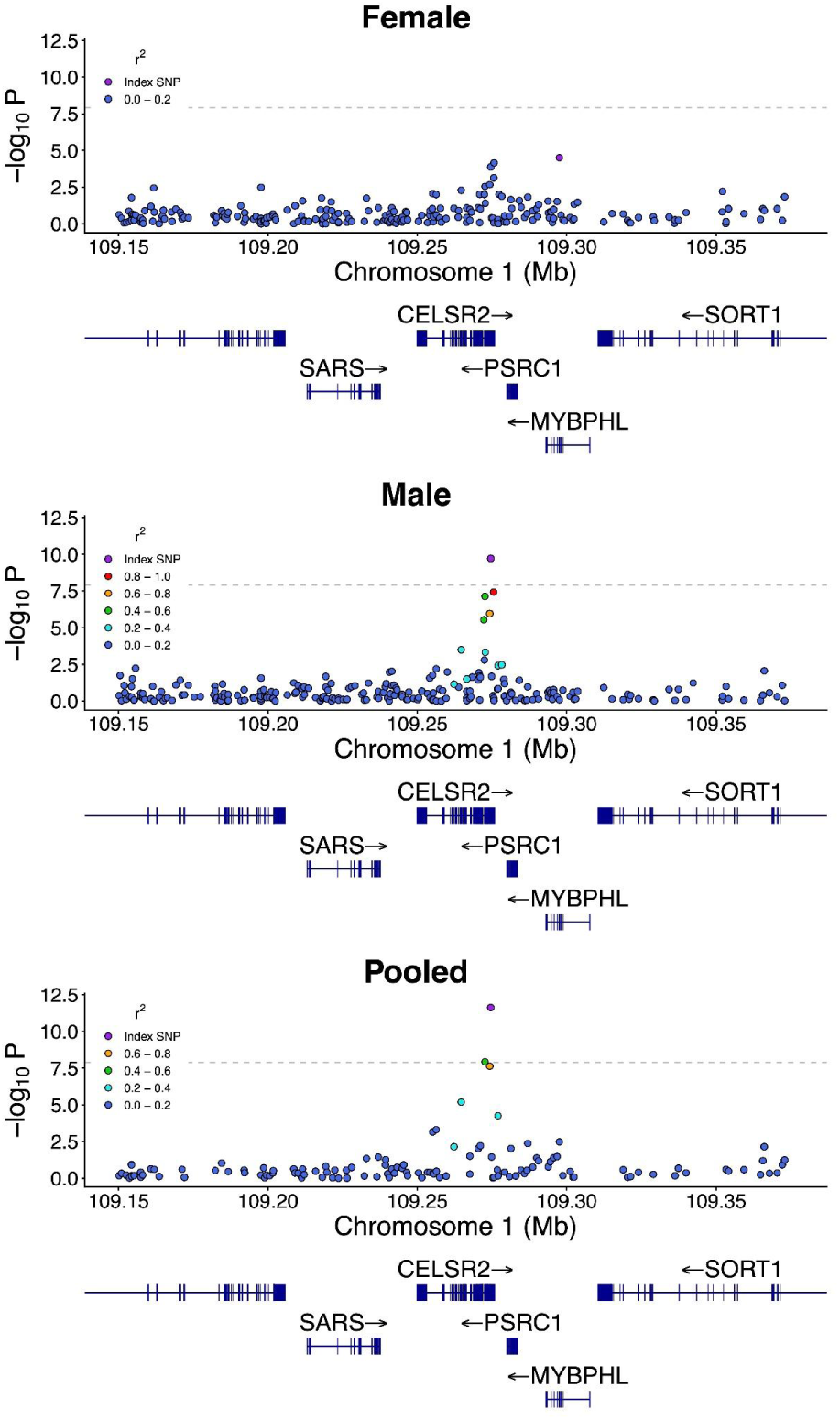
Regional association plots for *CELSR2* showing male-specific association with HDL-C.

**Figure 4B.**
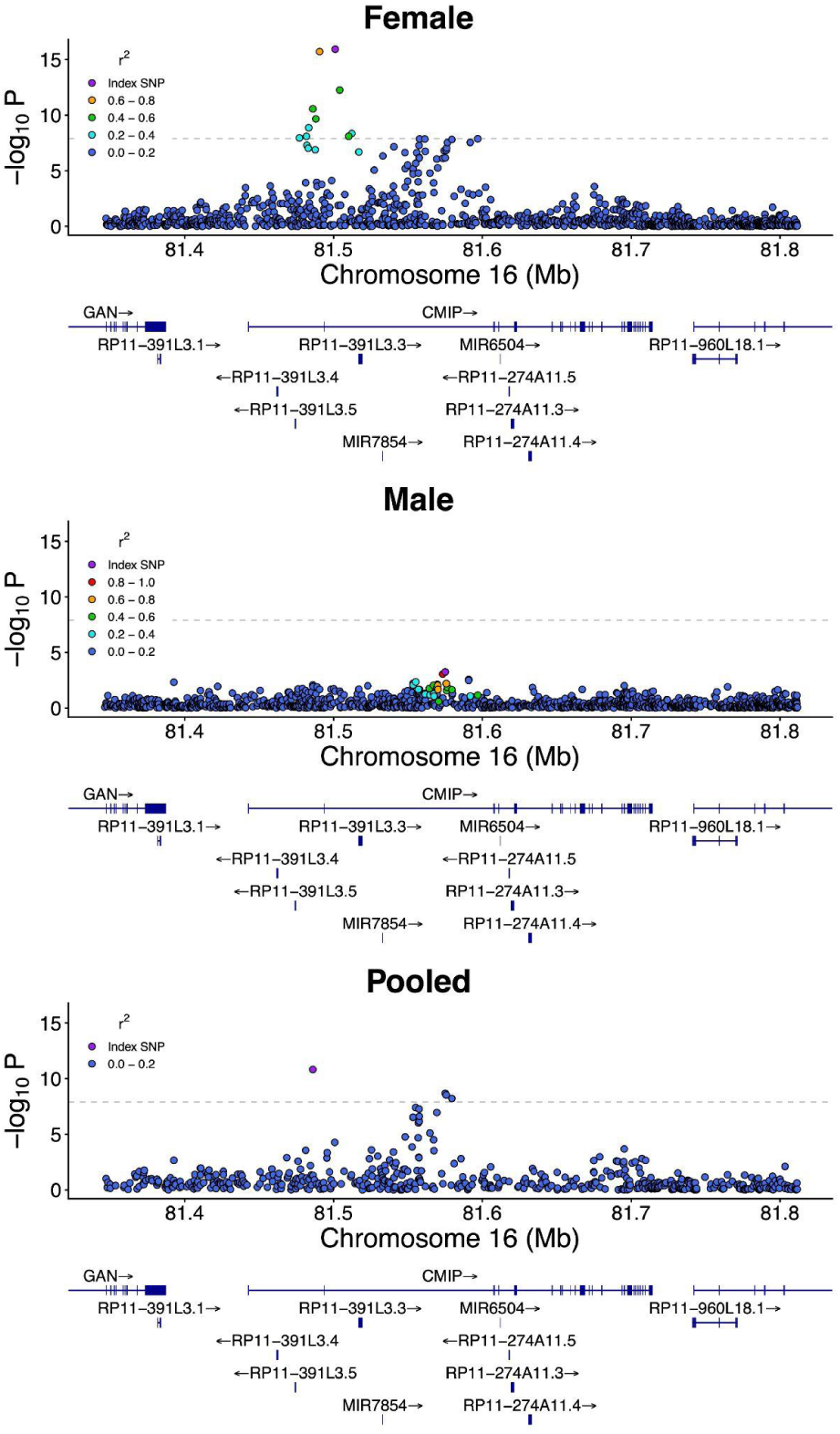
Regional association plots for *CMIP* showing female-specific association with TG.

**Figure 5.**
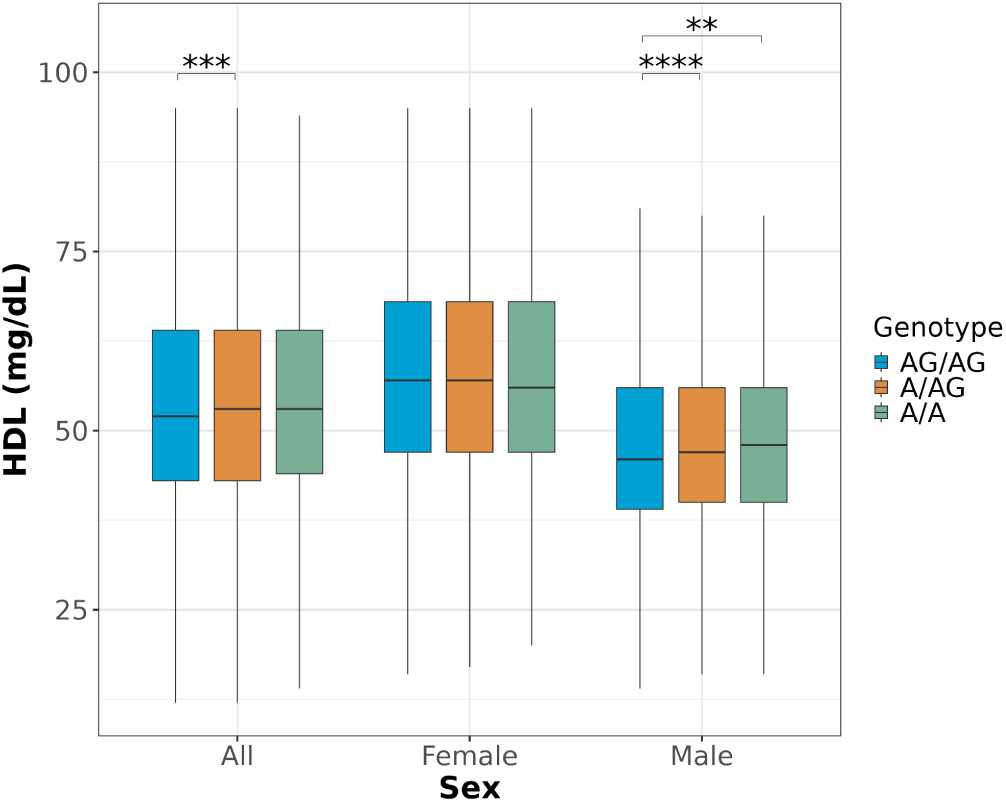
rs7528419 at chr1:109274570:A:G shows male-specific significance for HDL-C (CELSR2). Boxplots show lipid profile levels stratified by genotype and sex for variants exhibiting sex-specific associations.Values outside of 1.5 inter-quartile range from quartiles were not shown. Significance threshold are noted in asterisks using the following convention: ****: Tuckey adjusted p-value < 0.0001 ***: Tuckey adjusted p-value < 0.001 **: Tuckey adjusted p-value < 0.01 *: Tuckey adjusted p-value < 0.05

In addition, we have found *CMIP* to show female-specific significant association with TG (Figure 4B). *GPAM* and *PLTP* showed female-specific significant association with HDL-C (Supplementary Figure 2A). We observed female-specific dosage effects of increasing TG at chr16:81501185:A:G (rs2925979) for *CMIP* (Supplementary Figure 3D), and female-specific dosage effects of decreasing HDL-C at chr10:112150963:A:G (rs1129555) for *GPAM* (Supplementary Figure 3A) and at chr20:45916308:T:C (rs58847685) for *PLTP* (Supplementary Figure 3B). Interestingly, male participants carrying 1 alternative allele at chr20:45916308:T:C for *PLTP* gene showed statistically significant decrease in HDL-C, but no statistically significant difference was observed between male participants carrying 2 alternative alleles and 2 reference alleles at the loci (Supplementary Figure 3B). We have also found *ZPR1* to show female-specific significant association with LDL-C. We observed a statistically significant difference for female participants carrying 1 alternative allele at chr11:116778201:G:C (rs964184), but no dosage effect was observed at this variant (Supplementary Figure 3C).

## Discussion

We observed age-group- and sex-specific differences in lipid profile in AOU data. To investigate genetic control underlying such difference, we ran sex-specific GWAS and found sex-specific association analysis revealed novel sex-specific genetic risk loci for HDL-C, LDL-C, and TG that has not previously been reported to be sex-specifically related to lipid metabolism.

Previous research in age- and sex-specific lipid profiles has been conducted in a dutch cohort study [41]. While informative, the study was limited by the relatively homogeneous composition of the study population [41]. In our study, we observed consistent patterns of lipid variation between females and males across age groups. Leveraging the ancestry-diverse AOU easured differences in mean lipid levels using Cohen’s *d* to provide standardized effect estimates.

Although our study did not reveal the mechanisms behind sex specificity of these genes in their association to lipid profiles, future research can look into the potential mechanisms. Previous research has shown that *CELSR2–PSRC1–SORT1* gene cluster is associated with lipid levels and coronary heart disease [42]. *CELSR2* knockdown experiment in L02 human fetal hepatocyte cell line showed decreased lipid accumulation due to dysregulation of endoplasmic reticulum (ER) homeostasis, more specifically due to impairment in unfolded protein response [43]. Another mice experiment has indicated that sexual dimorphism before puberty in ER stress may be due to lack of testosterone in males [44]. We hypothesize that the male-specific association of *CELSR2* with HDL-C levels may be related to sex dimorphism in ER stress related to sex hormones. Future studies can investigate how male-specific regulation in ER stress is related to lipid metabolism.

The prevailing hypothesis for *CMIP*’s female-specific effects is an interaction with female hormones and fat distribution. An intronic *CMIP* polymorphism (rs2925979) has been associated with adiponectin levels, which is an insulin-sensitizing adipokine that raises HDL-C and lowers TG [45,46]. A previous meta-analysis of 114 studies involving 320,485 individuals showed *CMIP* to have association for waist-to-hip ratio (adjusted for BMI), indicating that *CMIP* may be involved in abdomen fat accumulation. On the other hand, another study shows that the *CMIP* risk allele (rs2925979*T) paradoxically correlates with lower BMI and waist circumference yet higher type 2 diabetes risk in females [47]. It is possible that in women the variant induces insulin resistance and dyslipidemia through pathways beyond generalized adiposity. In summary, we suspect that *CMIP* variants affect TG values in a sex-specific pattern through mediating adiponectin and shift in fat distribution. Further studies are warranted to provide more mechanistics behind such sex-specific effects.

Our study has several limitations including calculated LDL-C values. The first one is the dependent relationship between LDL-C values and other lipid measurements. While TC and HDL-C are directly measured from the serum, LDL-C is commonly calculated by the Friedewald equation, which is LDL-C = TC − (HDL-C) − (TG/5), which brings bias in itself and when LDL-C values are explored in GWAS [48]. Previous research has shown that calculated LDL-C values have higher variation and bias at lower LDL-C levels and advised against using LDL-C smaller than 70mg/dL for treatment decisions [49]. In GWAS for LDL-C values, genetic variants associated with other lipid types may also indirectly appear associated with LDL-C. In fact, in our analysis, *ZPR1* showed female-specific association with LDL-C, which was further validated by observation of statistically significant difference in LDL-C values for female participants with C/C (alt/alt) genotype compared to C/G (alt/ref), but not for males. *ZPR1* also demonstrated similar, despite weaker, female-specific association with TC. Future research can look deeper into sex-specific association of *ZPR1* with LDL-C after taking into consideration the calculative nature of many LDL-C measurements.

Another limitation of our study is the extensive missingness of statin use record. In our research cohort, 21,422 (17.1%) participants had statin usage history with start time within 1 year before the lipid measurement time in our study, among which 6,919 (32.3%) did not have statin use end time available. Extensive missingness in statin use end times, along with variable time it takes for statin to take effect in different individuals, make it not feasible to definitively determine if the LDL-C values used in study was affected by statin. Previous research in lipid GWAS using electronic health record or biobank data have handled repeated lipid measurements and lipid-lowering therapy in several ways. One study defined pre-treatment LDL-C to be the median of all LDL-C values before the first mention of any statin treatment [50]. Another biobank-level study on LDL-C adjusted values for statin use by dividing measured LDL-C by 0.7 to approximate pre-treatment levels [51]. However, such adjustment did not take into account interindividual variability of lipid lowering effect of statin [52]. Given the extent of missingness in statin end dates in our dataset and the resulting uncertainty about treatment status at any given blood draw, we opted instead to use the highest recorded LDL-C value across each participant’s lifetime electronic health record, reasoning that this “peak” LDL-C is most likely to precede initiation of lipid-lowering therapy and that the accompanying lipid panel is therefore less likely to be influenced by statins. Nevertheless, future research with access to more comprehensive statin use history can more thoroughly investigate genetic risk factors underlying hyperlipidemia while taking into consideration the effect of lipid lowering medications.

## Conclusion

In our research, we investigated lipid panel values across 124,920 participants in the AOU research program. We characterized sex-stratified HDL-C, LDL-C, TG, TC distributions across lifespan. Our findings revealed sex-specific gene associations with lipid profiles such as *CELSR2* (for male) and *CMIP* (for female). The study paves the way for further evaluation in mechanisms underlying sex-specific genetic associations with lipid profiles and provides foundation for sex-specific risk evaluation of cardiovascular risk.

## Data Availability

All data used in study are available for authorized users in All of Us researcher workbench.

## Acknowledgement

We gratefully acknowledge AOU participants for their contributions, without whom this research would not have been possible. We also thank the National Institutes of Health’s AOU Research Program for making available the participant data examined in this study.

## Supplementary Figures

**Supplementary Figure 1.** Manhattan plots for sex-specific and pooled GWAS. Manhattan plots display female-only, male-only, and pooled genome-wide association results for HDL-C, LDL-C, TG, and TC in the pooled cohort. Each point represents a genetic variant plotted by chromosomal position and –log₁₀(P). The horizontal dashed line denotes the genome-wide significance threshold after bonferroni correction.

**Supplementary Figure 1A.**
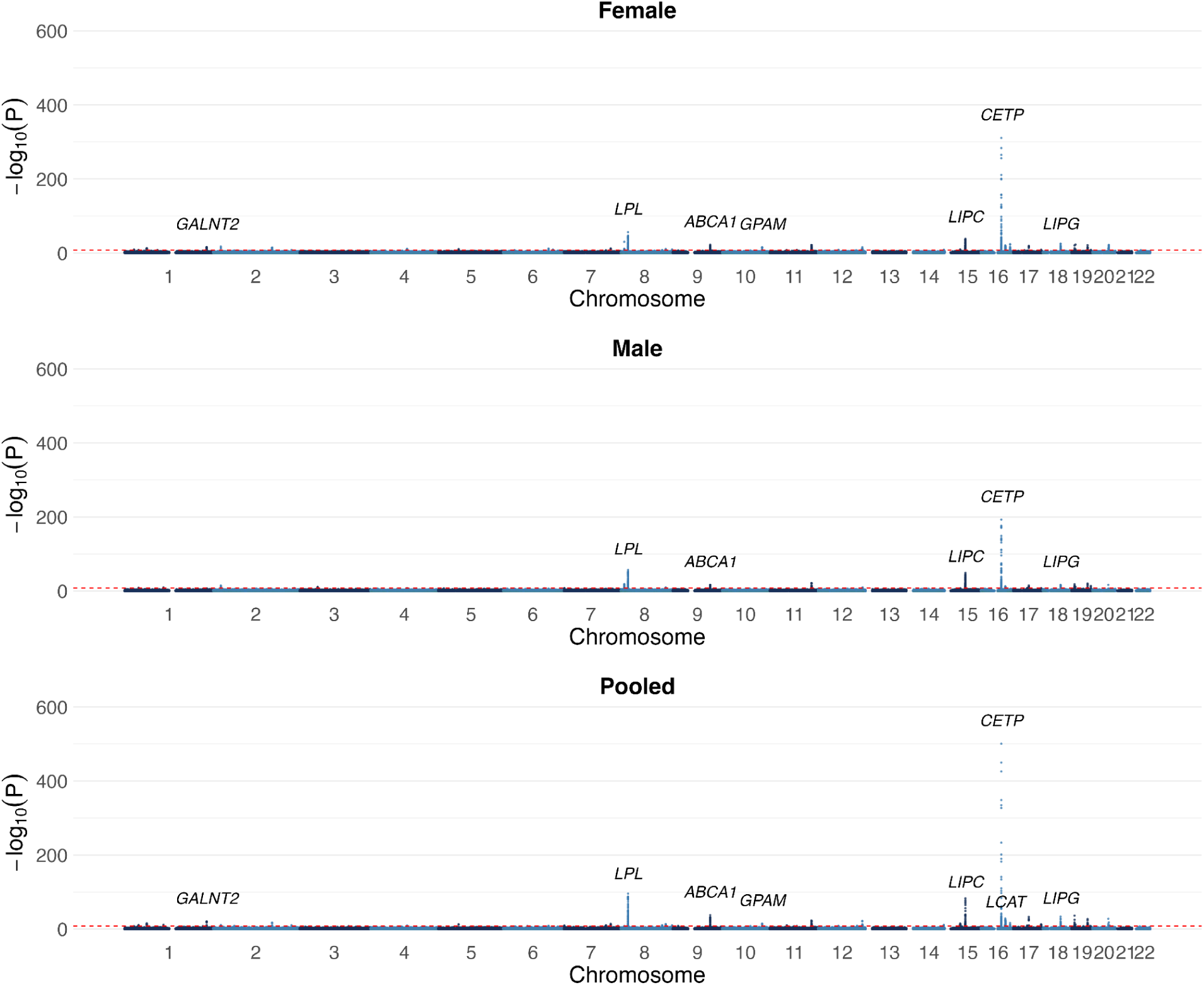
Manhattan plots for sex-specific and pooled GWAS for HDL-C.

**Supplementary Figure 1B.**
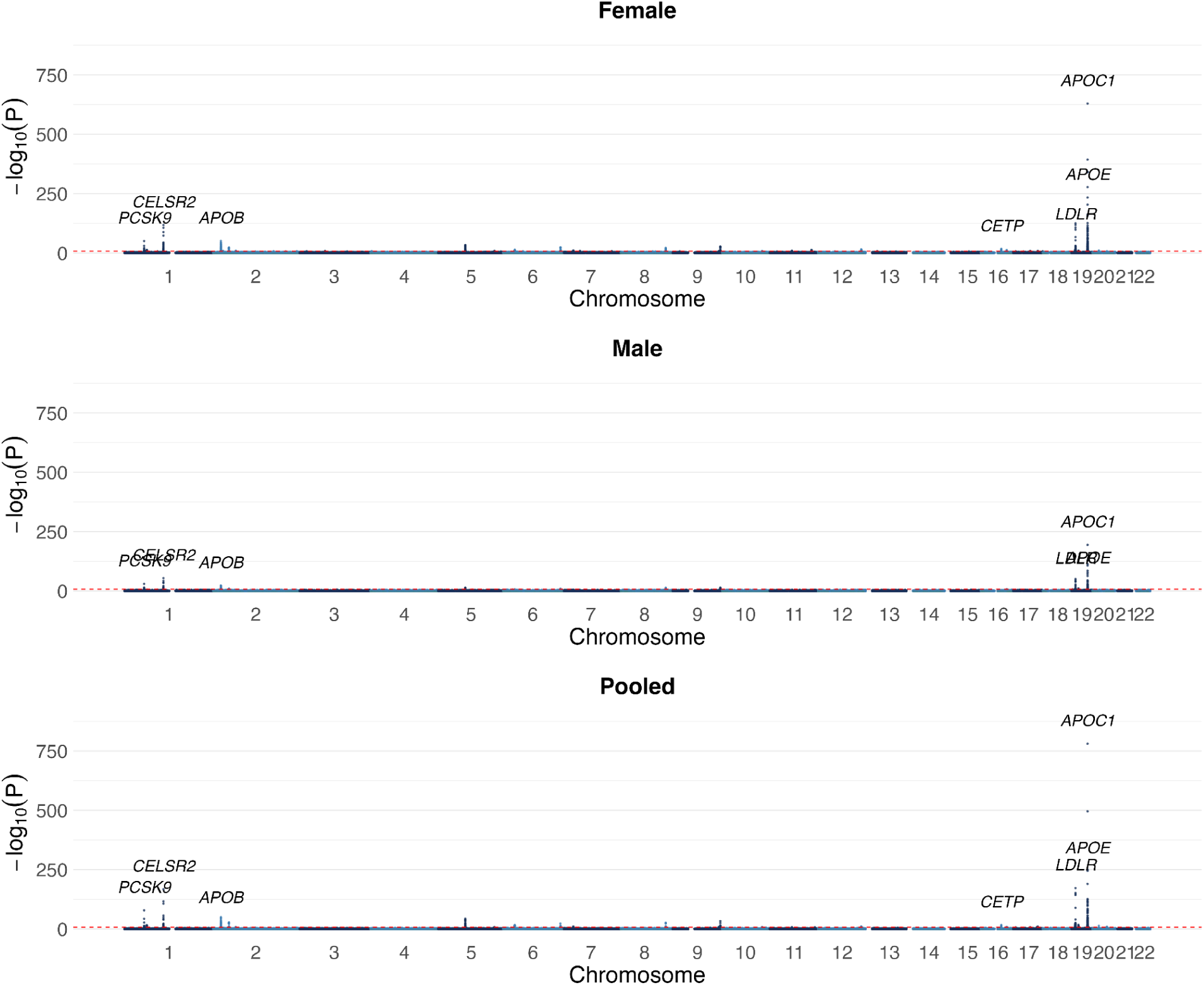
Manhattan plots for sex-specific and pooled GWAS for LDL-C.

**Supplementary Figure 1C.**
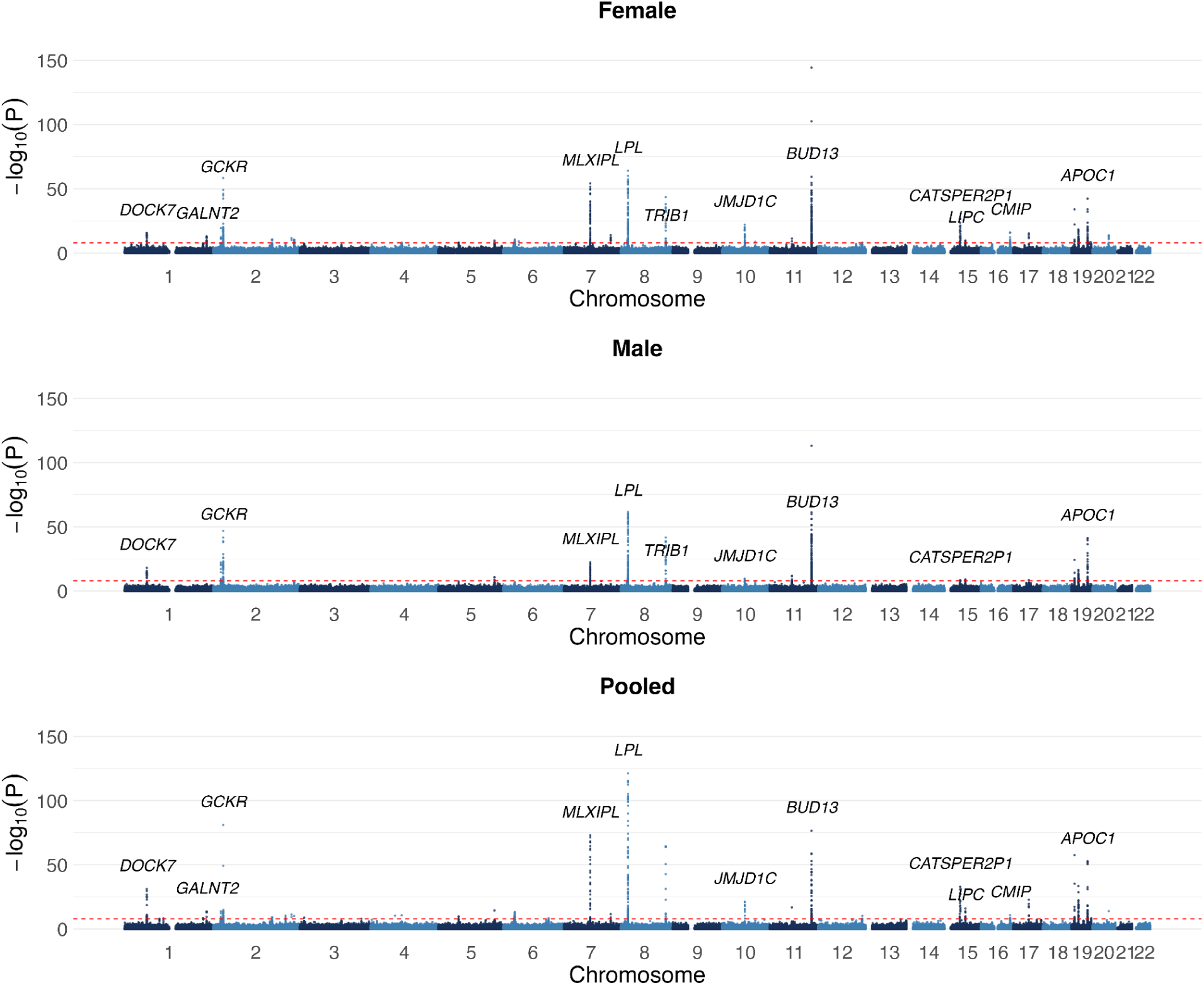
Manhattan plots for sex-specific and pooled GWAS for TG.

**Supplementary Figure 1D.**
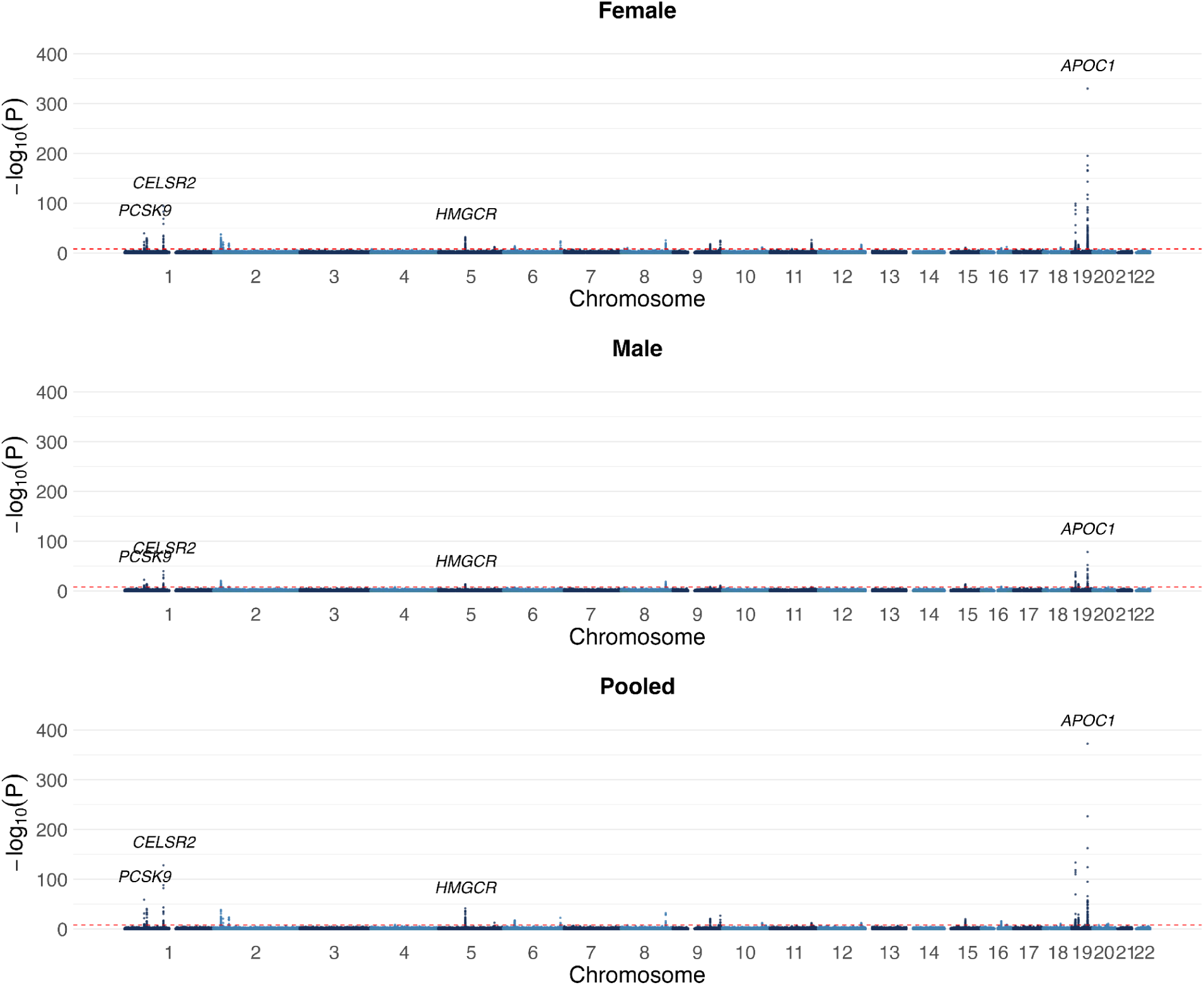
Manhattan plots for sex-specific and pooled GWAS for TC.

**Supplementary Figure 2.** Regional Association plots showing sex-specific associations. Regional association plots illustrate sex-specific associations. Variants are shown with –log₁₀(P) values plotted against genomic position, and are colored according to linkage disequilibrium (r²) with the lead variant. Gene models and transcript orientations are displayed below each plot to contextualize the association signals using the locus zoom package.

**Supplementary Figure 2A.**
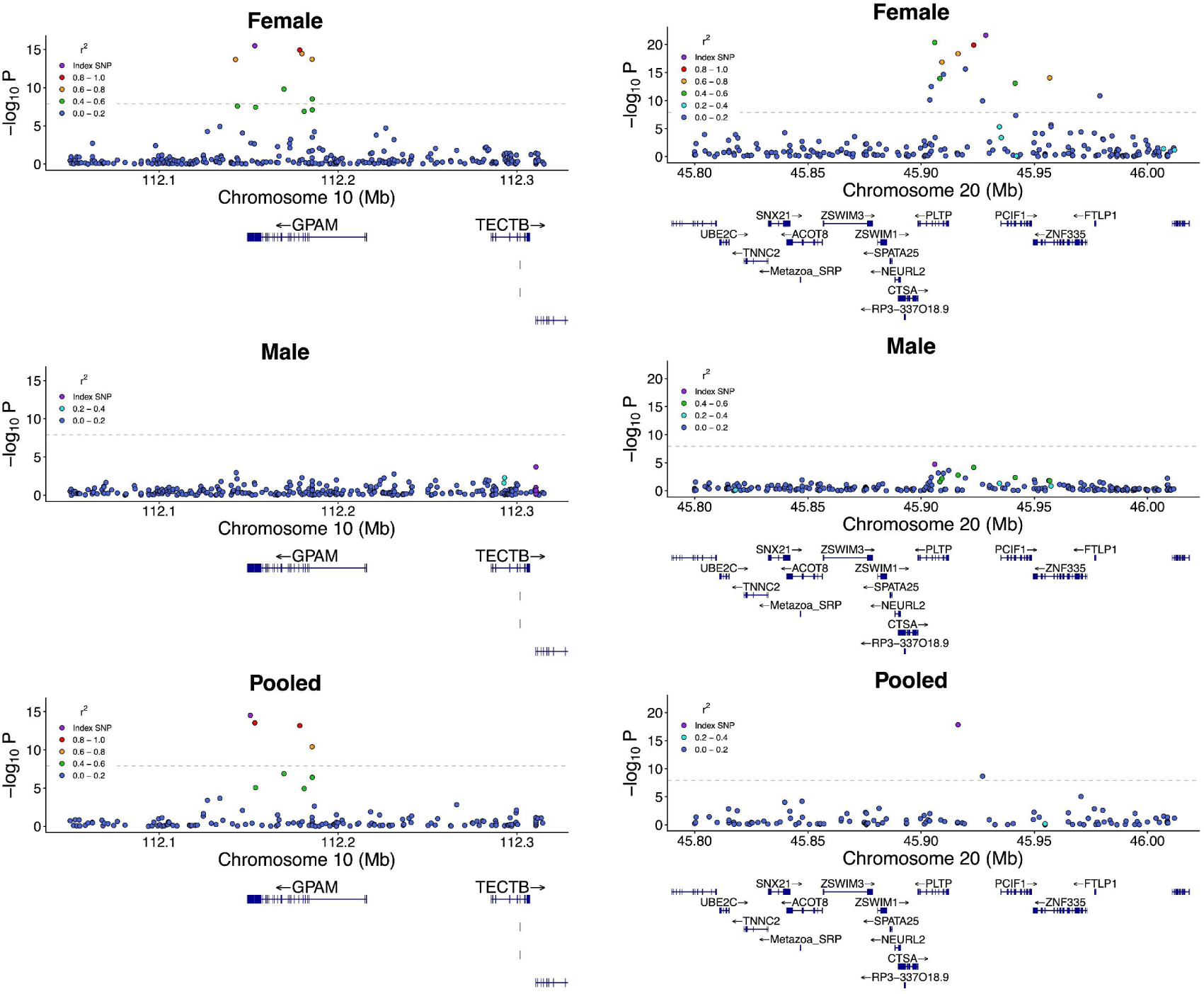
Regional association plots for *GPAM* (left) and PLTP (right) showing female-specific associations with HDL-C.

**Supplementary Figure 2B.**
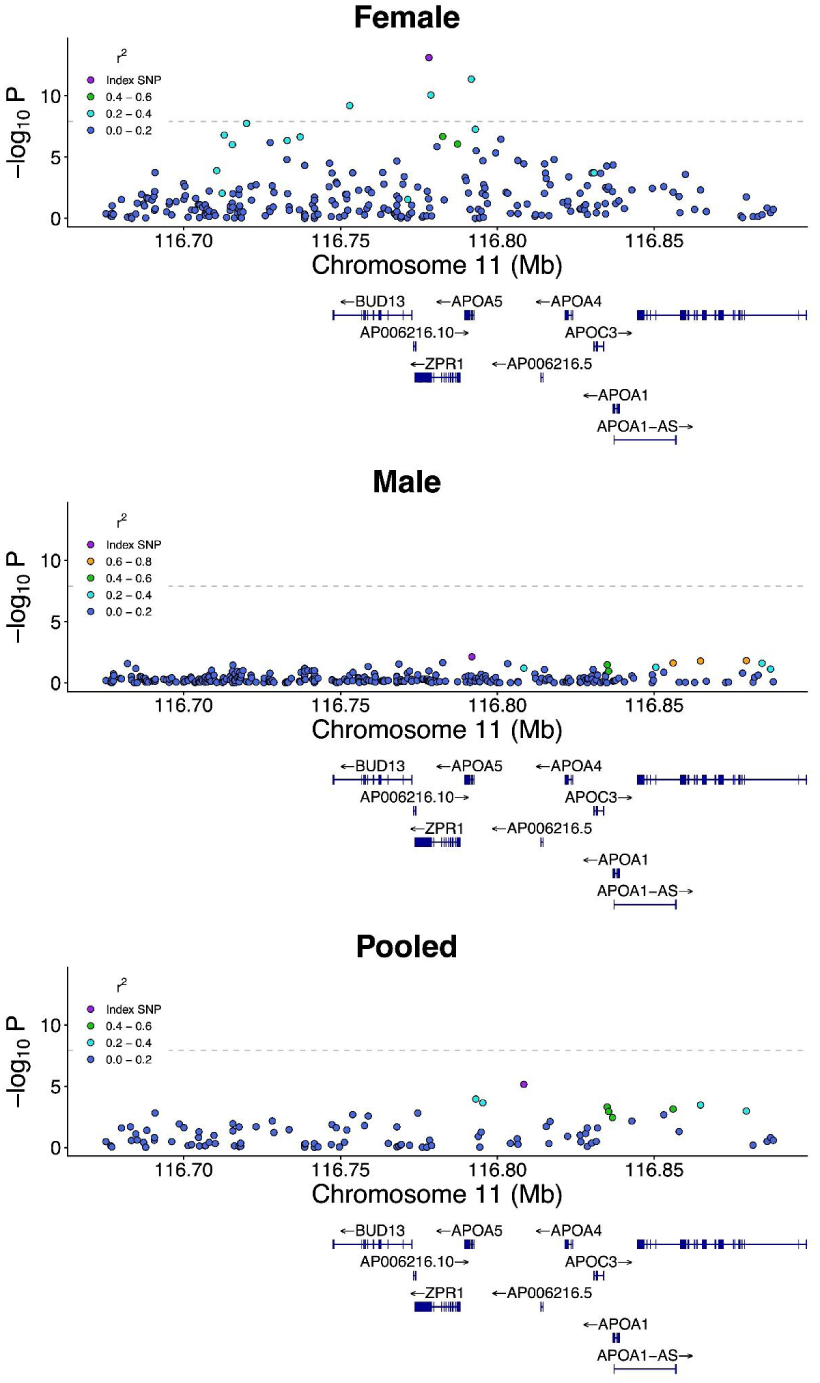
Regional association plots for *ZPR1* showing female-specific association with LDL-C.

**Supplementary Figure 3.** Lipid panel values for individuals with different genotypes at sex-specific loci. Boxplots show lipid profile levels stratified by genotype and sex for variants exhibiting sex-specific associations.Values outside of 1.5 inter-quartile range from quartiles were not shown. Significance threshold are noted in asterisks using the following convention: ****: Tuckey adjusted p-value < 0.0001 ***: Tuckey adjusted p-value < 0.001 **: Tuckey adjusted p-value < 0.01 *: Tuckey adjusted p-value < 0.05

**Supplementary Figure 3A.**
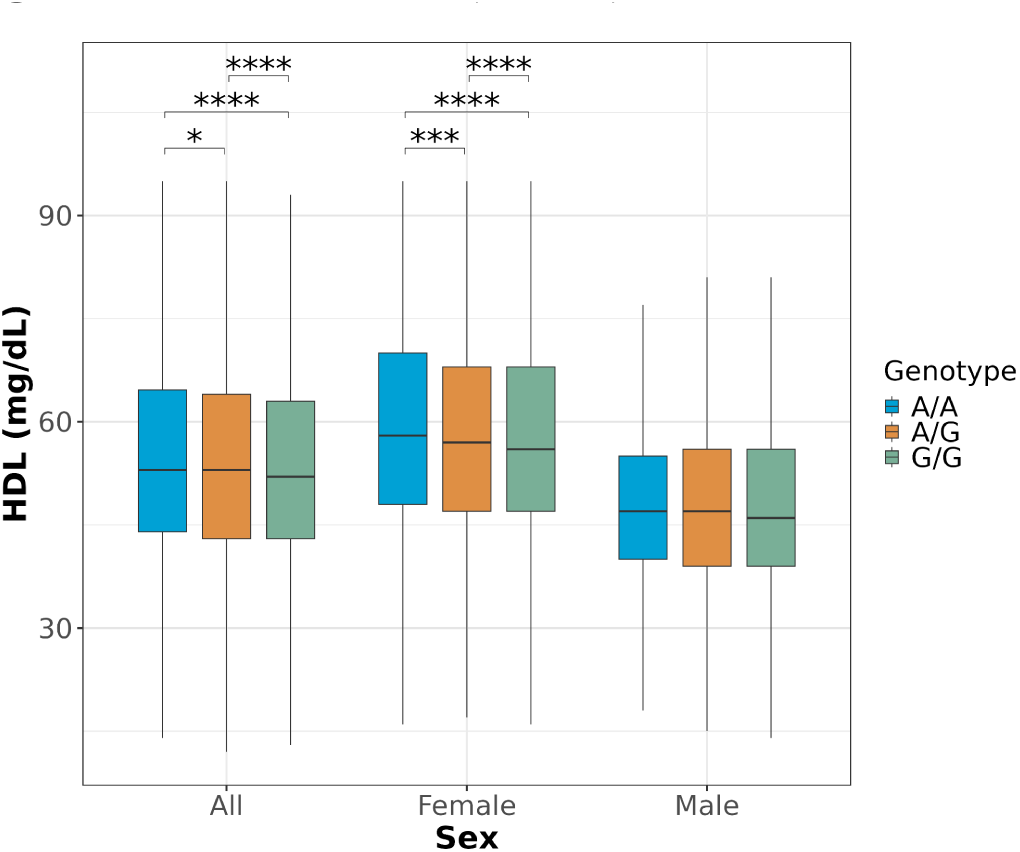
rs1129555 at chr10:112150963:A:G shows female-specific significance for HDL-C (*GPAM*).

**Supplementary Figure 3B.**
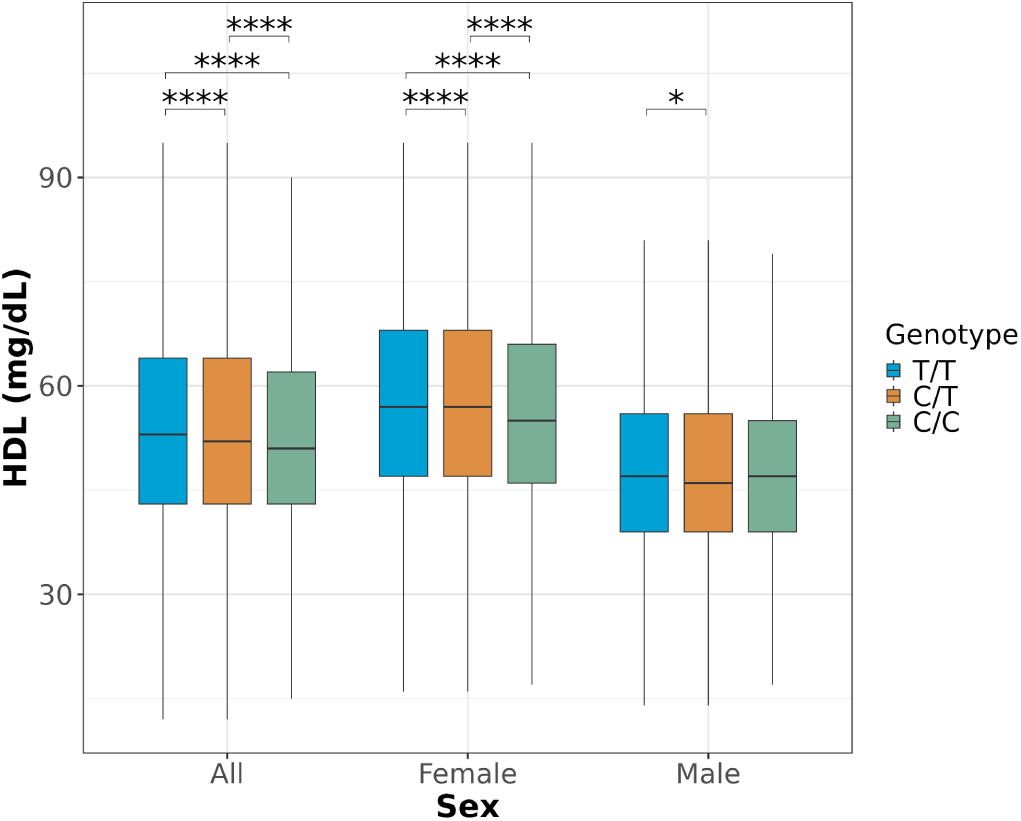
rs58847685 at chr20:45916308:T:C shows female-specific significance for HDL-C (*PLTP*).

**Supplementary Figure 3C.**
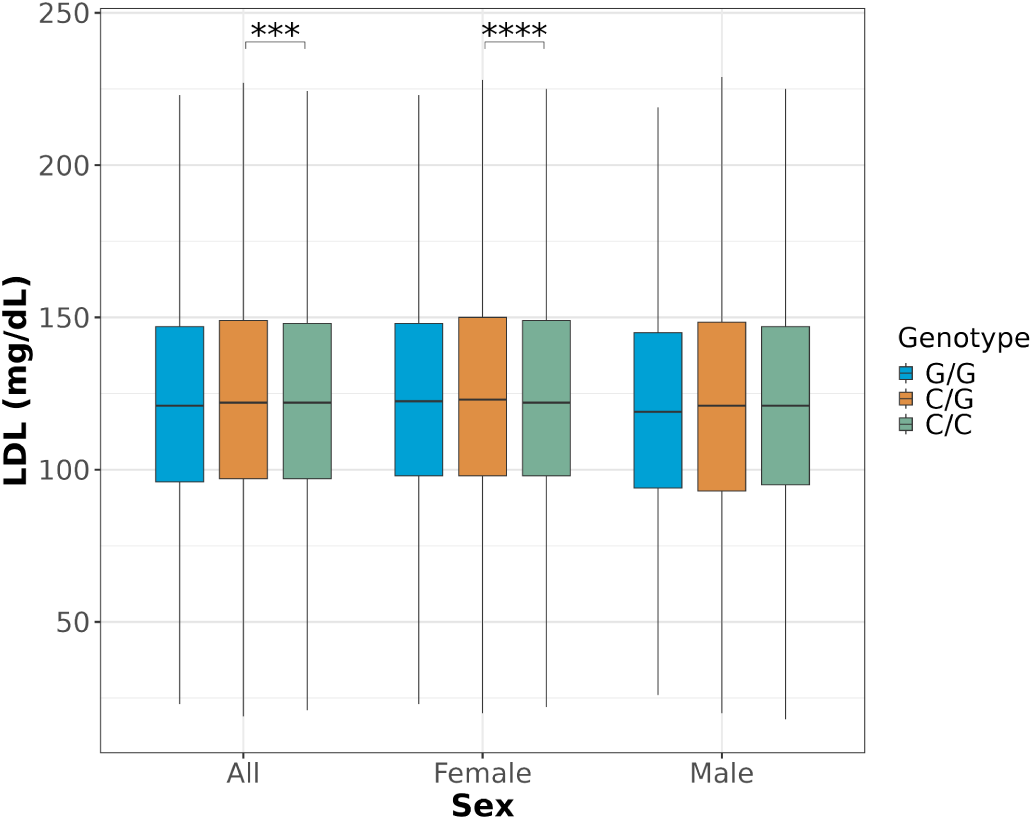
rs964184 at chr11:116778201:G:C shows female-specific significance for HDL-C (*ZPR1*).

**Supplementary Figure 3D.**
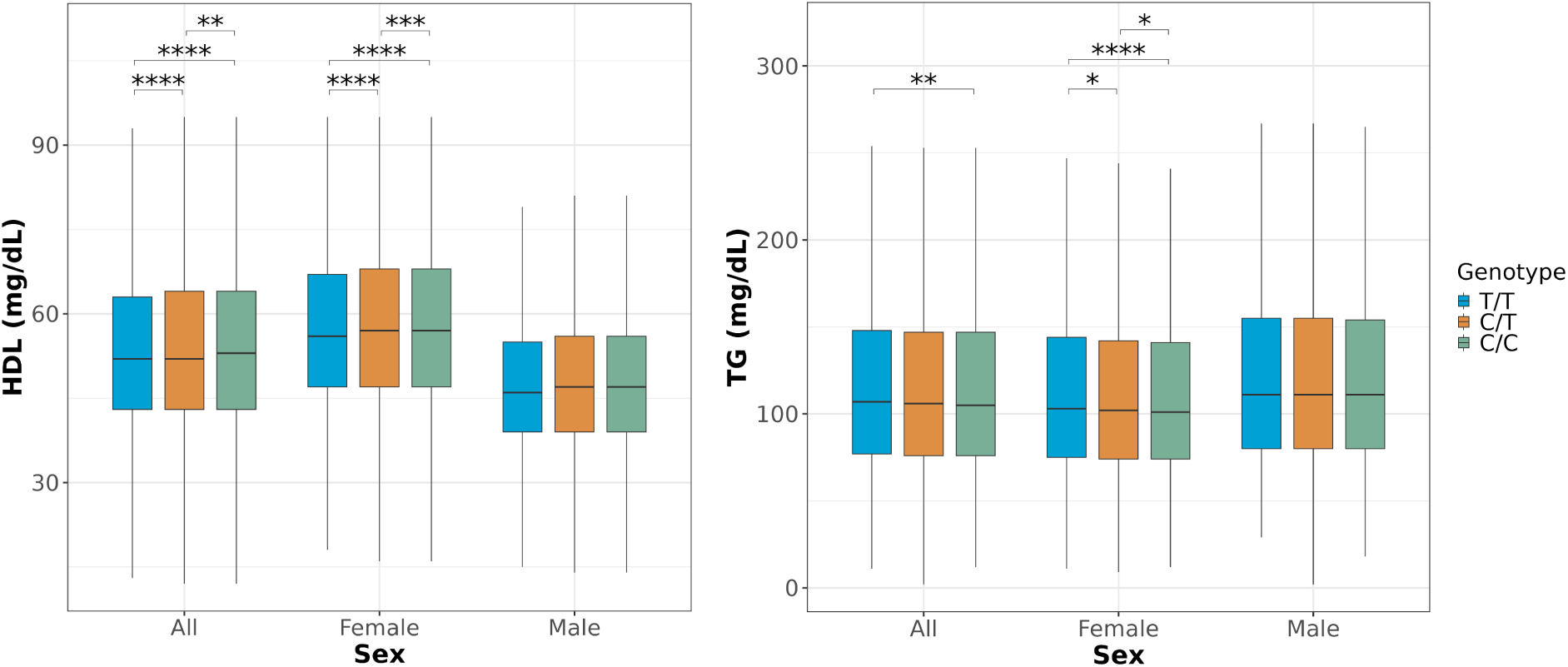
rs2925979 at chr16:81501185:A:G shows female-specific significance for HDL-C and TG (*CMIP*).

**Supplementary Figure 4.**
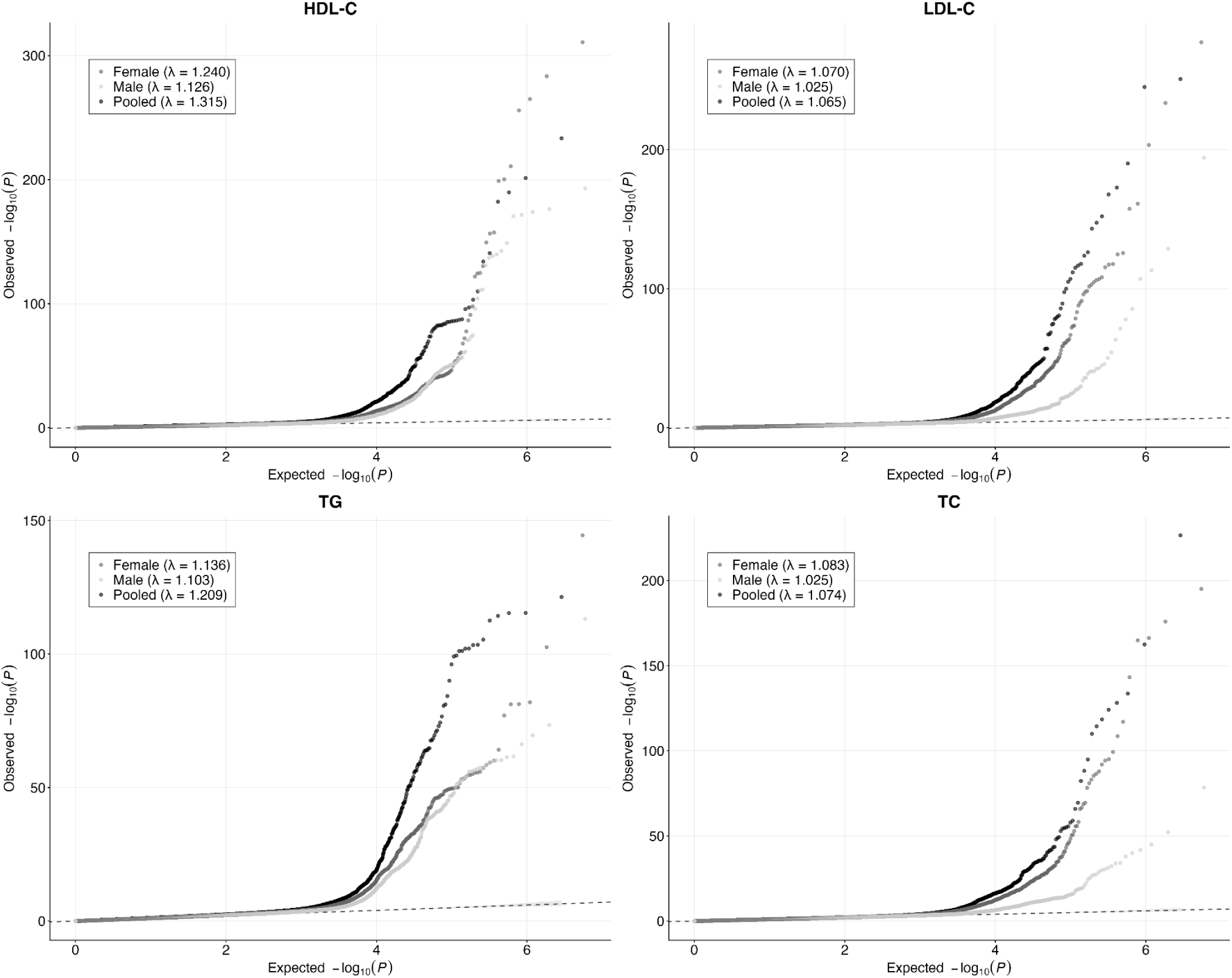
Quantile-quantile (QQ) plots for pooled and sex-specific GWAS for HDL-C, LDL-C, TG, and TC. For each trait, observed –log10(P) values from the pooled (all participants) GWAS and from sex-specific (female-only, male-only, and pooled) GWAS are plotted against the expected –log10(P) values under the null distribution. Lambda genomic inflation factors (λGC) for each analysis are indicated in the panels.

## Supplementary Tables

**Supplementary Table S1.** SNP level summary table of statistically significant variants of pooled GWAS run. Beta coefficients were estimated using the reference allele as the effect allele.

**Supplementary Table S2.** Gene level summary table of statistically significant genes of pooled GWAS run.

**Supplementary Table S3.** SNP level summary table of statistically significant variants of female-specific GWAS run. Beta coefficients were estimated using the reference allele as the effect allele.

**Supplementary Table S4.** SNP level summary table of statistically significant variants of male-specific GWAS run. Beta coefficients were estimated using the reference allele as the effect allele.

**Supplementary Table S5.** Gene level summary table of statistically significant variants of female-specific GWAS run.

**Supplementary Table S6.** Gene level summary table of statistically significant variants of male-specific GWAS run.

